# Evaluation of Primary Prescription Nonadherence Post-discharge from an Inpatient Psychiatry Unit at a Large Urban Community Hospital – an Observational Study

**DOI:** 10.1101/2021.09.02.21262083

**Authors:** Meghana Ganjam, Ysatis Ruiz, Luis G. Allen, Rosemary Persaud

## Abstract

**Introduction:** Here, we aimed to evaluate primary prescription nonadherence post-discharge from an acute inpatient psychiatric unit. Prescription nonadherence is a concern across all patient care settings, with primary nonadherence defined as not picking up prescribed medication from the pharmacy or not delivering prescriptions to the pharmacy. Secondary nonadherence, defined as filling a prescription but not taking the medication as prescribed, was not tracked in this study. The incidence of nonadherence can vary widely across settings and has been reported to range from 3– 86%. This is a particular concern in patients with a primary psychiatric diagnosis both in an outpatient and inpatient setting.

**Design:** The prescription fill rate of eligible patients was tracked on days 7 and 21 post-discharge from acute inpatient psychiatric units of an acute care multispecialty urban community teaching hospital.

**Results:** In total, 72 patients aged 18 and above (44%, women) were analyzed. A primary nonadherence incidence of 43% was found, which did not vary significantly across the analyzed variables of age, sex, or primary diagnosis.

**Conclusion:** Primary nonadherence is a significant issue in this population. Strategies, such as the implementation of “med-to-bed” programs and use of longer acting injectables when appropriate, would help in increasing adherence. Further research, including the evaluation of other variables that affect nonadherence, is needed to identify and develop steps to overcome the obstacles to adherence.

## 1 Introduction

The treatment of psychiatric health conditions is a global health challenge that contributes to 30% of the nonfatal disease burden (1), and mental health problems, such as depression, represent a growing and increasingly important public health concern. Approximately 20.6% (51.5 million) of adults aged 18 and over in the US suffer from mental illness in any given year, and approximately 25.5% (13.1 million) of these patients suffer from severe mental illness (2). Neuropsychiatric disorders are a leading cause of disability, accounting for 18.7% of years of life lost to disability and premature mortality (3); further, evidence suggests that disorders, such as posttraumatic stress disorder (PTSD) and depression, increase the incidence of heart disease and dementia among other medical conditions (4, 5, 6). The management of psychiatric conditions as well as of the associated physical maladies is made more difficult by the increased incidence of the nonadherence to medication (7) in individuals with a mental health diagnosis. The World Health Organization (WHO) has defined nonadherence as “a case in which a person’s behavior in taking medication does not correspond with agreed recommendations.” The high-risk group in this category of patients comprises of the subset of those who were sick enough to require inpatient psychiatric hospitalization and were then discharged. There have been few studies that were focused on evaluating primary nonadherence in this patient population. These individuals are particularly vulnerable, and nonadherence leads to poor outcomes, including readmission along with an elevated risk of other comorbid conditions (8, 9). The aim of our study was to gauge the incidence of primary nonadherence by evaluating the failure to fill prescriptions for medication within 21 days of discharge from an inpatient psychiatric unit of a large urban community hospital.

## 2 Materials and Methods

### 2.1 Study Design

The study population included 73 inpatients from the Central Florida region who were discharged between January 2017 and May 2017 from a psychiatric unit. The study was determined not to require institutional review board oversight. Eligible patients had an overnight or longer stay at either of the two inpatient psychiatric units at the hospital: a 20-bed capacity unit catering to a more elderly or medically complex population and a 24-bed capacity unit catering to a younger and typically less medically complex population. Patients were transferred between these units based on their medical needs and on hospital staffing and were not tracked separately. Eligible patients were discharged with electronic prescriptions for the required medications, which were sent to a pharmacy of the patient’s choice. Patients who were transferred to another medical unit or who were discharged to a structured living facility, such as an assisted living facility or a skilled nursing facility, were excluded. Patients who did not have a pharmacy of choice and who received handwritten or printed prescriptions instead of electronic prescriptions were also excluded. In total, 73 subjects qualified for inclusion over a period of 120 days. The prescription status of one of the subjects could not be verified, and therefore, this subject was excluded from the study.

### 2.2 Data Collection

Pharmacies where the prescriptions were sent were contacted on days 7 and 21 to confirm if prescriptions were picked up. Based on whether the medications were picked up, the prescriptions were coded using binary points (0, not filled and 1, filled); similarly, the medication pickup status was coded for both days 7 and 21 (0, no and 1, yes).

Medications that were prescribed for “as needed” (PRN) use were not considered. Over-the-counter medications (such as folic acid and nicotine replacement products) were not considered. Claims-related data and communication with the pharmacy were used to verify whether a refill was not needed for continuing the prescription (refill too early). The dataset did not contain any insurance information. The primary diagnosis of “acute psychosis” was changed to psychosis. Incidental notes were made regarding several patients who were uninsured or underinsured and who did not use insurance to pay for prescriptions. This was addressed in the study by calling pharmacies to verify pickup or noncompliance without having to rely on insurance claims made for prescriptions.

### 2.3 Variables

The collected patient information included sex, age, and primary diagnosis. No insurance information was collected. Each variable was tested separately because of the small sample size.

### 2.4 Data Analyses

All tests were two-tailed, and a value of 0.05 was set for statistical significance. All tests were performed using IBM SPSS version 21 software (IBM, Armonk, NY, USA).

## 3 Results

The sex distribution of the study population is presented in **Table 1**. The study population included approximately 44% of women and 56% of men. The diagnosed psychiatric disorders in the study population are listed in **Table 2**.

**Table 1.** Sex distribution of the study population

|  |  | Frequency | Percent | Cumulative percent |
| --- | --- | --- | --- | --- |
| Valid | Female | 32 | 43.8 | 43.8 |
|  | Male | 41 | 56.2 | 100.0 |
|  | Total | 73 | 100.0 |  |

**Table 2.**
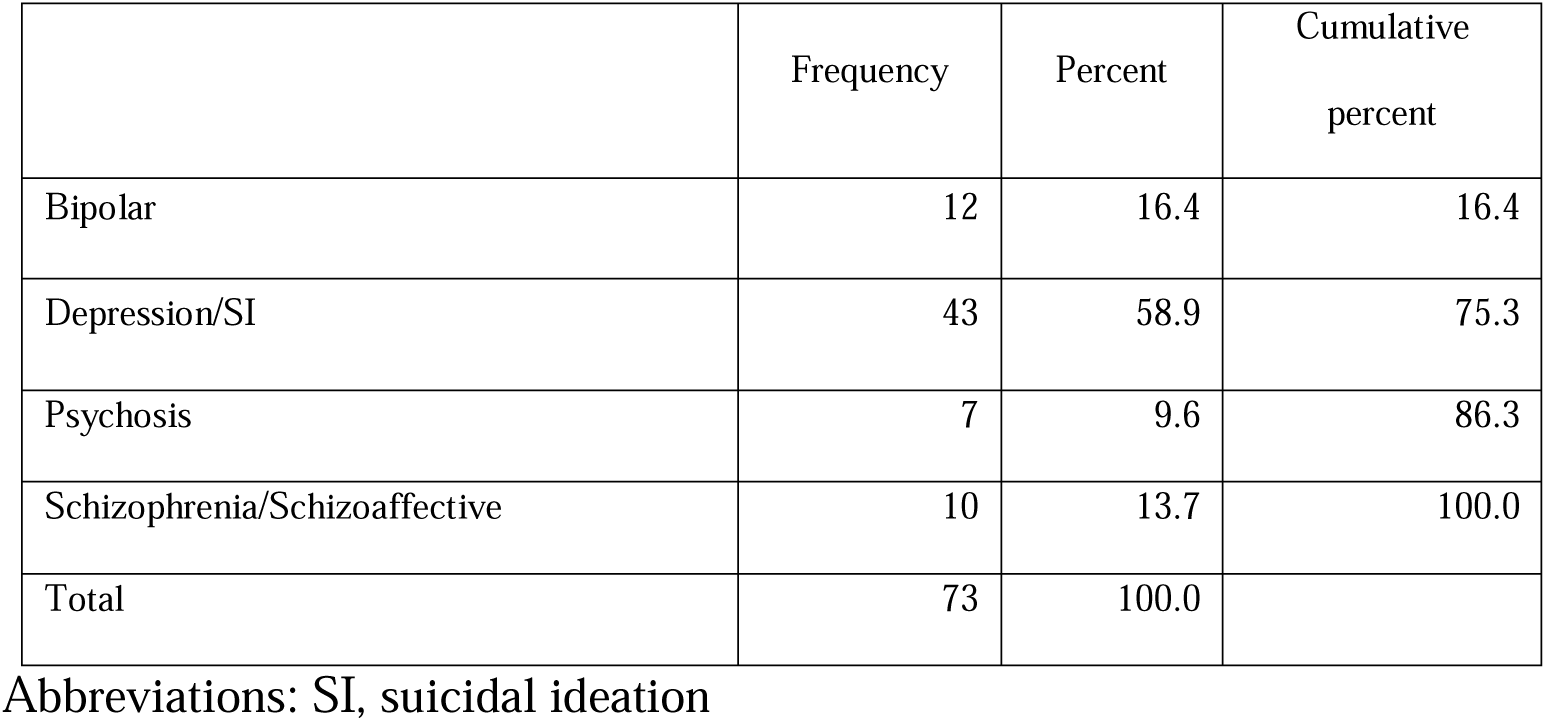
Description and frequency of common psychiatric disorders in the study population

|  | Frequency | Percent | Cumulative percent |
| --- | --- | --- | --- |
| Bipolar | 12 | 16.4 | 16.4 |
| Depression/SI | 43 | 58.9 | 75.3 |
| Psychosis | 7 | 9.6 | 86.3 |
| Schizophrenia/Schizoaffective | 10 | 13.7 | 100.0 |
| Total | 73 | 100.0 |  |

Abbreviations: SI, suicidal ideation

The study population included a large proportion of individuals diagnosed with depression (∼60%). The adherence to prescription pickup on days 7 and 21 is presented in **Tables 3A and 3B**. Of the 72 subjects, 35 (49%) picked up their prescriptions by day 7 (95% confidence interval, 37%– 60%). By day 21, 41 subjects (57%) were noted to have picked up their prescriptions (95% confidence interval, 45%–68%). There was no statistically significant difference in the rate of pickup between men and women (p = 0.394) based on the results of a chi-square test of independence (**Table 4**). Similarly, age was not associated with the rate of pickup, as there was no statistical difference in the age of patients according to whether they picked up their prescriptions (p = 0.620) based on an independent t-test for means (**Table 5**). There was no statistical difference in the rate of pickup by primary diagnosis (p = 0.235) based on a chi-square test of independence (**Table 6**).

**Table 3A.**
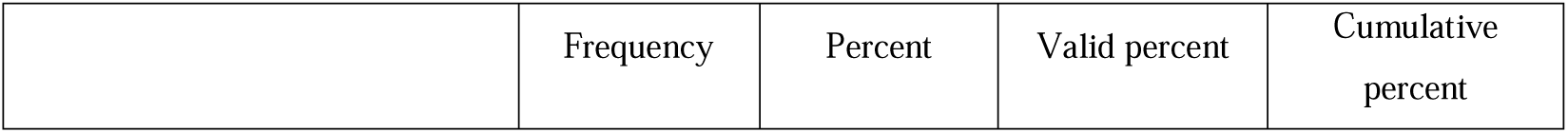
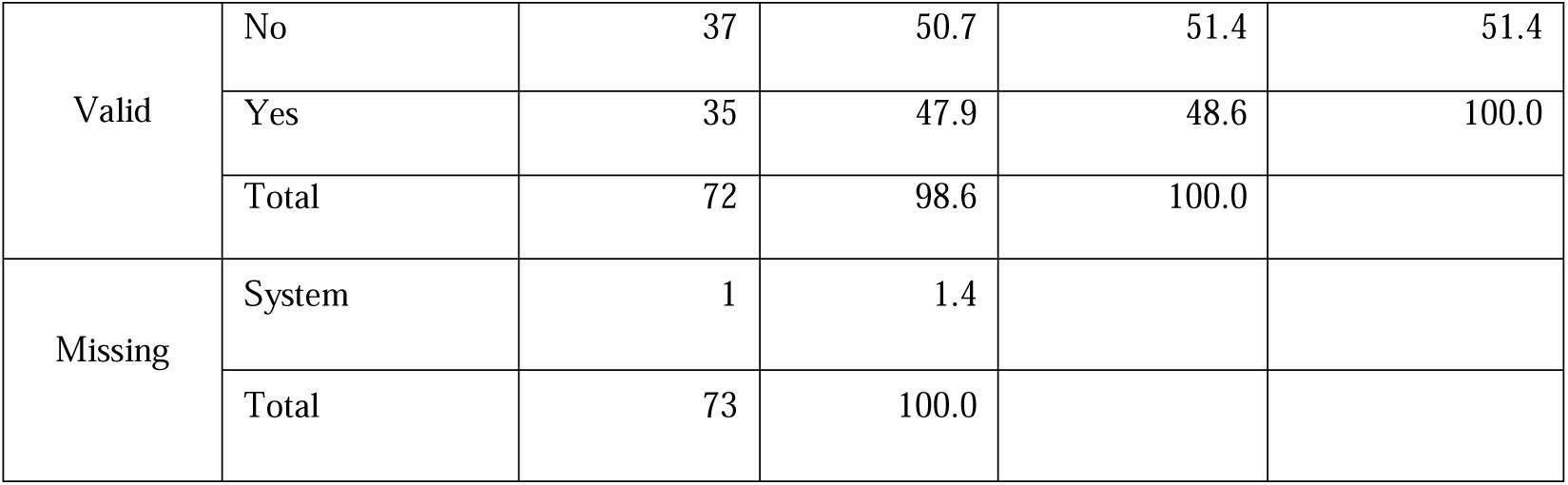
Adherence to prescription pickup by day 7

**Table 3B.**
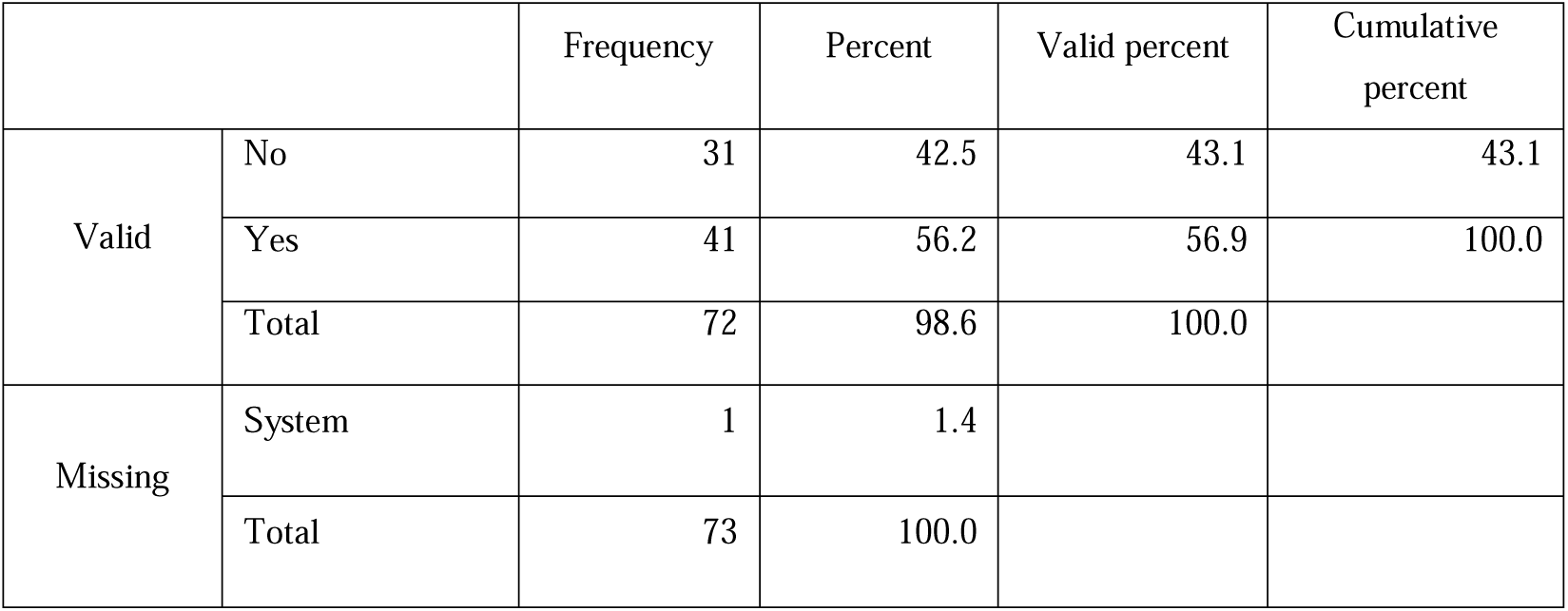
Adherence to prescription pickup by day 21

**Table 4.**
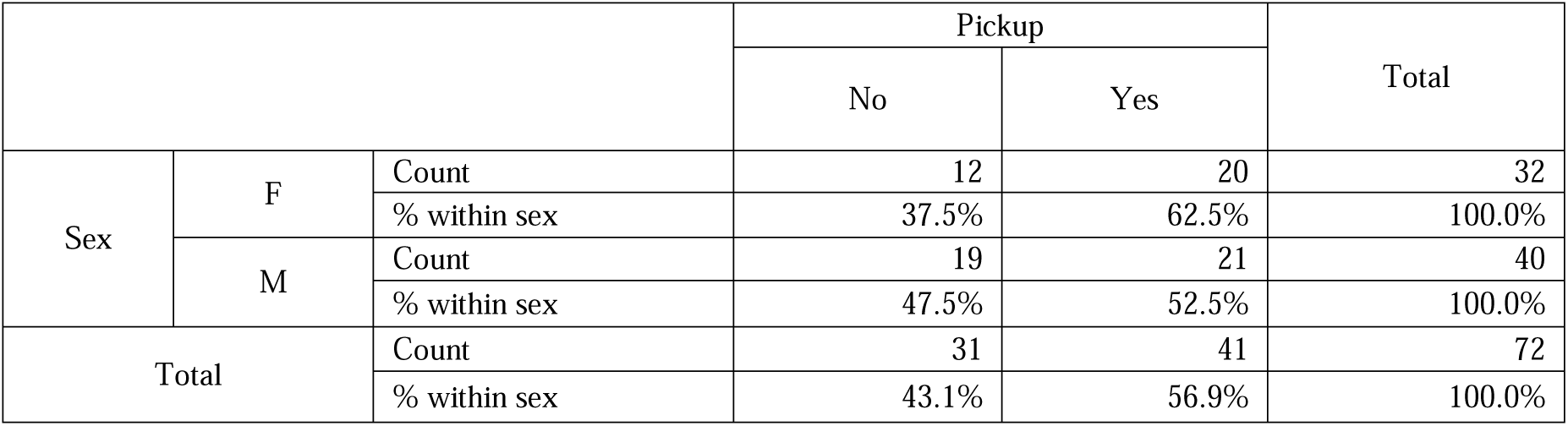
Association between sex and prescription pickup

**Table 5.** Association between age and prescription pickup

|  | Pickup | N | Mean | Std. deviation | Std. error mean |
| --- | --- | --- | --- | --- | --- |
| Age | No | 31 | 50.84 | 9.832 | 1.766 |
|  | Yes | 41 | 49.41 | 13.439 | 2.099 |

Abbreviations: Std., standard

**Table 6.** Association between primary diagnosis and prescription pickup

| Primary diagnosis |  |  | Pickup |  | Total |
| --- | --- | --- | --- | --- | --- |
|  |  |  | No | Yes |  |
|  | Bipolar | Count | 6 | 6 | 12 |
|  |  | % within sex | 50.0% | 50.0% | 100.0% |
|  | Depression/SI | Count | 14 | 28 | 42 |
|  |  | % within sex | 33.3% | 66.7% | 100.0% |
|  | Psychosis | Count | 5 | 3 | 8 |
|  |  | % within sex | 62.5% | 37.5% | 100.0% |
|  | Schizophrenia/schizoaffective | Count | 6 | 4 | 10 |
|  |  | % within sex | 60.0% | 40.0% | 100.0% |
| Total |  | Count | 31 | 41 | 72 |
|  |  | % within sex | 43.1% | 56.9% | 100.0% |
| Chi-square test |  | Value | df | Asymp. Sig. (2-sided) |  |
| Pearson chi-square |  | 4.260<sup>a</sup> | 3 | .235 |  |
| Likelihood ratio |  | 4.272 | 3 | .234 |  |
| No. of valid cases |  | 72 |  |  |  |

Abbreviations: SI, suicidal ideation

## 4 Discussion

Prescription medication nonadherence, a major public health concern that is prevalent across different healthcare settings globally, leads to poor outcomes and increased healthcare costs (10). In a 2003 report on medication adherence, the WHO noted that “increasing the effectiveness of adherence interventions may have a far greater impact on the health of the population than any improvement in specific medical treatments” (11). In the current study, the observed incidence of primary nonadherence to prescription medications was 43% based on the behavior of not picking up prescriptions 21 days post-discharge. There is a lack of standardization of the appropriate time period between prescription generation and pickup for the measurement of primary nonadherence (12,13). For the purpose of this study, we chose two timepoints, days 7 and 21. Unfilled prescriptions are returned to stock by day 7 at most pharmacies, and the longest period is usually less than 21 days. The differences in primary nonadherence based on the variables that were assessed (age, sex, or primary diagnosis) were not found to be statistically significant. In some prior studies, it was found that primary nonadherence was higher among patients with severe mental illness, such as schizophrenia (14), but this was not the case in this study population. Multiple factors affect nonadherence, including patient beliefs regarding medication efficacy and side effects, ease of accessing the medication, complexity of the medication regimen, socioeconomic status, insurance status, and marital status among others (15,16). Among patients with chronic illness, it is estimated that nearly 50% do not take medications as prescribed (11). This adversely affects disease outcomes across a wide spectrum of chronic diseases, including chronic obstructive pulmonary disorder, cardiovascular disease, hypertension, and diabetes. Nonadherence becomes particularly important in the subset of patients recently discharged from an acute care setting post-stabilization (17, 9). Within this subset of patients, there are limited data on primary nonadherence post-discharge from acute psychiatric facilities in the US. There are several existing obstacles that hinder the accurate tracking of these prescriptions. Fragmented healthcare systems and payers and the slow adaption to electronic medical records and subsequently to electronic prescription among psychiatric hospitals make prescriptions more difficult to track, as do the larger geographic area that psychiatric hospitals usually serve as compared to acute care multispecialty hospitals. Nonadherence in the subset of patients discharged following hospitalization in a psychiatric unit has been shown to lead to worse outcomes, higher costs, and increased readmissions (7, 9, 18, 19). The goal of this study was to determine the incidence of nonadherence in this specific patient population in order to enable healthcare facilities to develop strategies for improving adherence in this vulnerable patient population. Some of the strategies may include a universal “med-to-bed” (20) program, in which all patients receive a supply of the needed medications along with medication-related education at discharge. Other interventions include communication between the hospital physician and patient’s primary care physician prior to discharge and early post-discharge outpatient follow-up appointments with primary care physicians and psychiatrists (11), which would enable the earlier identification of nonadherence. The use of equivalent long-acting injectable formulations (21), with the initial dose administered prior to discharge, is yet another strategy.

In 2020, the US spent more than 225 billion dollars on mental health. There are over 170,000 residents occupying inpatient and other 24-h residential treatment beds on any given night, which is an average of over 53.6 patients per a population of 100,000 (22). This number does not include patients incarcerated for mental illness. Along with the decrease in the number of available inpatient psychiatric beds over the past several decades, the average length of stay in patients admitted to inpatient acute psychiatric facilities has also decreased from several weeks to ∼10 days with most patients staying for shorter durations (23). Individuals with mental health diagnoses account for a disproportionate sum of the total health expenditure even though only a small proportion of this is spent on mental health (24). The 30-day unplanned readmission rate among patients discharged from inpatient psychiatric hospitals is approximately 20% (25).

This study has some limitations. Only patients who chose a predesignated pharmacy and who agreed to have their prescriptions sent electronically were included for the ease of tracking. It is unclear whether the inclusion and tracking of other patients would have altered the results. The primary admitting diagnosis alone was used to stratify the patients, and it is unclear if the addition of secondary diagnoses would have altered the results. Insurance data were not accurately collected and were excluded from the study. This could have provided additional factors related to nonadherence. Improving medication adherence is a key path toward improving healthcare, and promoting primary adherence is the first essential step. This study that has assessed primary nonadherence in this important population subset is of value as our results could aid in gauging the severity of this problem and in formulating strategies for improving the delivery of effective healthcare. Improvements in the care of this population will not only improve mental healthcare but will also decrease the overall healthcare expenditure. The hospital where this study was conducted has since implemented a med-to-bed program in order to more effectively serve this patient population.

## Data Availability

The data that support the findings of this study are available from the corresponding authors (MG and RP) upon reasonable requestNo

## 6 Author Contributions

MG - Review of Data, Research and Analysis, References, Documentation and Write up

YR - Formatting and References

LGA - Supervising Physician

RP - Design and Implementation

## 1 Data Availability Statement

The data that support the findings of this study are available from the corresponding authors (MG and RP) upon reasonable request.

